# Participant outcomes evaluation of the DWELL (Diabetes and WELLbeing) type 2 diabetes 12-week psychoeducational self-management programme across four European countries

**DOI:** 10.1101/2025.01.13.25320460

**Authors:** Eleni Hatzidimitriadou, Sharon Manship, Rachael Morris, Thomas Thompson, Julia Moore, Sabina Hulbert, David Vernon

## Abstract

**Introduction:** Type 2 diabetes mellitus (T2DM) is a lifelong condition that has large societal, economic and clinical implications, and treatment should be supported by healthy lifestyle factors. Interventions for effective self-management are essential to the sustainability of treatment, however there is no standard approach.

**Research Design and Methods:** Six hundred and five participants diagnosed with Type 2 Diabetes Mellitus were recruited from four countries (UK, France, Netherlands, Belgium) to complete the 12-week DWELL (Diabetes and WELLbeing) psychoeducational intervention. The programme was delivered at community and hospital-based settings and comprised of four key areas: education, nutrition, physical activity and wellbeing. Metabolic health (weight, waist size, BMI and HbA1c) and self-reported psychological measures were taken at four points: pre- and post-intervention, and two follow up points (at 6 and 12 months) to assess the impact of the programme.

**Results:** Participants showed a significant reduction in all metabolic health measures, with improvements in both weight and BMI being maintained at 6-month follow-up. Participation in the programme also led to enhanced levels of participant empowerment, with significant improvements also seen in perceptions of diabetes, eating behaviours, mental and physical health, and self-care behaviours.

**Conclusions:** The study results demonstrated that an empowerment-based, holistic and flexible approach to diabetes self-management education programmes has a wider impact in improving longer term coping behaviours which help in achieving and sustaining positive metabolic and psychological changes.

**Key Messages:** 

**What is already known on this topic:** Diabetes education has evolved from a compliance and knowledge-oriented approach to an empowerment and self-management-oriented approach. Yet, type 2 diabetes self-management education (DSME) programmes are mainly evaluated in relation to impact on metabolic (glycaemic) outcomes than on wider psychosocial outcomes.

**What this study adds:** Participation in the DWELL programme led to significant improvements in metabolic health measures and produced significant positive changes across a range of psychological measures such as patient empowerment, illness perceptions, eating behaviours and self-care behaviours.

**How this study might affect research, practice or policy:** The DWELL DSME programme which was co-designed with patients, healthcare professionals and family carers, focussed on patient empowerment and self-control, by offering flexibility and choice of options as well as peer support. Programme outcomes indicated that this approach led to positive changes in empowering and enabling health behaviour changes and improvements in metabolic health. The study adds to the body of knowledge of patient-led DSME practice with a holistic approach. Further research could shed light on the cultural and intersectional aspects of such approach that can inform more targeted programmes supporting T2DM patients with multiple co-morbidities such as serious mental health conditions.

## Introduction

Diabetes is one the top 10 leading causes of death and disability globally, with 1.5 million deaths attributed to the condition each year and a leading cause for blindness, kidney failure, heart attacks, stroke and lower limb amputation worldwide (WHO, online). The most common is Type 2 diabetes mellitus (T2DM) and in the past three decades its prevalence has risen dramatically in all income level countries, with ninety per cent having onset usually in later life (WHO online; Sun et al, 2022). Prevalence of T2DM has escalated more rapidly in low- and middle-income countries (LMICs) compared with high-income countries (HICs), with an estimated 79.4% of the global T2DM population residing in LMICs (IDF, 2021. In 2021, the estimated global annual cost of diabetes treatment was 966 billion USD (IDF, 2021), imposing a substantial health and economic burden on individuals, their families, and healthcare systems (Seuring, Archangelidi & Suhrcke, 2015).This rise is driven by a complex interplay of socio-economic, demographic, environmental and genetic factors, including urbanisation, an ageing population, decreasing levels of physical activity and increasing prevalence of overweight and obesity (IDF, online).

Type 2 diabetes mellitus (T2DM) has been identified as one of the most challenging chronic conditions to manage. Since the management of diabetes is mainly accomplished by patients and families, self-management has become the mainstay of diabetes care. Systematic reviews of research evidence show that consistent, careful self-management is supporting patients to achieve best outcomes (Ernawati, Wihastuti & Utami, 2021). Diabetes self-management education (DSME) is an intervention associated with improved glycated haemoglobin (HbA1c) and quality of life (QOL) and is recommended for all individuals with type 2 diabetes. Captieux et al (2018) conducted a meta-review of quantitative systematic reviews on supported self-management for people with type 2 diabetes. They included 41 systematic reviews incorporating data from 459 randomised controlled trials in diverse socio-economic and ethnic communities across 33 countries. All but one of the reviews confirmed HbA1c improvements of between 0.2% and 0.6% (2.2 – 6.5 mmol/mol) at 6-months post intervention but this change attenuated at 12 and 24 months. Impact on secondary outcomes was inconsistent and generally non-significant. Effective programmes tended to be multi-component and provide adequate contact time (>10 hours) (Captieux et al, 2018). Similarly, a recent review by Chowdhury et al (2024) has examined the effects of DSME interventions on glycaemic control, cardiometabolic risk, self-management behaviours, and psychosocial well-being among T2DM across LMICs. A total of 44 studies (n = 11838) from 21 LMICs were included in the review and analysis showed that DSME effectively reduced HbA1c levels by 0.64% (95% CI: 0.45% to 0.83%) and 1.27% (95% CI: -0.63% to 3.17%) for RCTs and quasi-experimental design studies, respectively. Also, findings showed an improvement in cardiometabolic risk reduction, diabetes self-management behaviours, and psychosocial well-being.

Despite the widely reported positive effect of DSME interventions on lifestyle changes and self-care of T2DM patients, there is currently no standard approach for diabetes self-management. Diabetes education has evolved from a compliance and knowledge-oriented approach to an empowerment and self-management-oriented approach (Hermanns et al, 2020). Diabetes Literacy, a pan-European project, which ran from 2012 to 2015, was a novel approach that aimed at strengthening patient diabetes self-management by exploring what exists and works (http://diabetesliteracy.com). The study surveyed the state of DSME in the 28 European Union Member States (EU MS) (n=249) and contrasted it with 3 non-EU countries with comparable Human Development Index score: Israel, Taiwan, and the USA (ITU) (n=130). Findings suggested that access to DSME varied greatly in EU MS - an average of 29% (range 21% to 50%) of respondents reported DSME programmes were tailored to people with limited literacy, educational attainment, and language skills versus 63% in ITU. More than half of adult T2DM patients and children/adolescents participated in DSME in EU MS as opposed to ITU. Prioritization of DSME (6.1 ± 2.8 out of 10) and the level of satisfaction with the current state of DSME (5.0 ± 2.4 out of 10) were comparable in the EU MS and ITU (Riemenschneider et al, 2018). The project team concluded that variation of DSME in Europe, partly due to different health systems, poses a great challenge for developing an EU-wide strategy towards these interventions.

In terms of DSME participation outcomes, evidence to date suggests that diabetes self-management education seems to have a greater impact on metabolic (glycaemic) outcomes than on mental health outcomes, but the latter are rarely assessed in the evaluation of these interventions, therefore there are knowledge gaps regarding the efficacy of diabetes self-management education on mental health (Hermanns et al, 2020). Indeed, evidence suggests that effective diabetes education and patient empowerment can lead to increased control over the condition and improved self-care behaviours (Wilson, 2021). Key aspects of improved health behaviours in relation to long-term conditions are perceived locus of control (LOC) and self-control, significant change determinants in self-management behaviours for people with T2DM which are not properly assessed in evaluation of such interventions (Macaden and Clarke, 2010; Nugent et al, 2015). Lack of internal LOC and poor self-control are linked to poor diabetes care behaviours, such as fears associated with glucose monitoring; lack of self-control over dietary habits; memory failure; and perceived lack of personal control, or external, such as support from family, peers, and health care providers. Greater internal locus of control and higher self-control are associated with improved physical health and psychological wellbeing, outcomes, which helps people to achieve their longer-term goals (Botha and Dahmann, 2023).

Nonetheless, a significant proportion of people with this life-long condition fail to engage in effective self-management at early stages of diagnosis and treatment, especially those in pre-diabetic phase or with aggravating comorbidities (e.g. severe mental illness) and socioeconomic factors (e.g. multiple deprivation) (Captieux et al, 2018). Ribu et al (2019) suggested a theoretical explanation for the daily life problems and challenges by those living with type 2 diabetes, proposing that the struggle to self‐manage and maintain new habits can be more or less difficult depending on the patient’s perceived conditions. They identified three situations: one where there is less struggle to let go of old habits, a second where there is more of a struggle to balance between what individuals want to do and what they ought to do and a third where they are giving up struggling. Study findings showed that healthcare professionals must employ flexible approaches to accommodate the heterogeneity of their patients’ situations, especially those who struggle the most, and adopt tailored approaches with varied options to meet their needs.

Issues that impact on an individual’s ability to self-manage their condition are multifactorial, including education, communication with healthcare providers, personal circumstances, provider issues and support (Abu et al, 2019; Nam et al, 2011; Wilkinson, Whitehead & Ritchie, 2014). Initiatives to increase effective, low-cost self-management are essential to the sustainability of treatment, such as education programmes that allow for incremental knowledge gain and experiential and vicarious learning and the provision of culturally sensitive care (Wilkinson, Whitehead & Ritchie, 2014). Cadzow et al (2014) suggest that community-level programmes have the potential to affect empowerment and self-efficacy and may be helpful in overcoming common barriers to self-management.

In order to address these wide-ranging issues, the Diabetes and WELLbeing (‘DWELL’) project aimed to develop, deliver and evaluate a 12-week psychoeducational programme for people with T2DM to empower them better self-manage the condition. The programme was co-developed by a diverse team of people with T2DM, their carers, healthcare professionals and academics (diabetic nurses, psychologists, counsellors, dieticians, fitness experts, and medical professionals). For the development and evaluation of the programme, we adopted the Medical Research Council framework for complex interventions (Craig et al, 2008) to enable comprehensive assessment of outcomes, processes and impact of the delivery of the programme.

In this paper, we report on quantitative participant outcomes’ evaluation results. Key research questions were:

1. What was the impact of the DWELL programme in terms of metabolic health, quality of life, levels of physical activity and self-care for T2DM patients?
2. What was the impact of the programme on self-management of diabetes in terms of participant attitudes and behaviours?

## Research Design and Methods

### Study design and ethical approval

A quasi-experimental, non-randomised, pre-post intervention design adopted a longitudinal mixed methods approach. Pre-post measures were taken at multiple time points; baseline (T0), end-of-programme (T1), at 6-months (T2) and 12 months (T3) follow up. Quantitative data was collected using the ‘DWELL Tool’, a set of validated questionnaires chosen to answer specific research questions, as well as an assessment form that included demographic, biomedical and metabolic data. Qualitative data was collected via end-of-programme focus groups, semi-structured interviews and researcher field notes. However, only quantitative data from pre, post and the initial follow-up (i.e., T2) are presented in this paper.

Ethical approval was obtained for the study from all programme sites, either via national ethics committees or organisational management approval.

### Participants

The main inclusion criteria were that participants be aged 18 years and over and had received a diagnosis of Type 2 Diabetes Mellitus (T2DM). At the outset 605 participants were recruited to the study from four countries (UK=299, France=203, Netherlands=55, Belgium=48). Ages ranged from 25 years to 93 years (Mean 62.5y, SD 10.0y), with 280 (47.2%) identifying as male and 313 (52.8%) as female. In terms of ethnicity 539 (92%) identified as *White*, 21 (3.5%) as *Asian*, 15 (2.5%) as *Black/African/Caribbean*, 8 (1.3%) as *Mixed* and 3 (0.5%) as *Other*. It should be noted that sample sizes at each time point may vary due to participants omitting to respond, possible attrition rates and exclusion criteria.

### Intervention

Participants were able to self-refer, or were referred by GPs, hospitals or healthcare professionals involved in their care to the 12-week DWELL programme. Full details of the 12-week DWELL programme and its practical organization can be found in Vanbosseghem et al. (2020). The programme comprised four key areas: education, nutrition, physical activity and wellbeing. As well as *core sessions* in each of the four areas, participants were signposted to further ‘pick and mix options’ that they could undertake outside of the programme, for example, local gym sessions or yoga classes. The programme was underpinned by peer support and self-management theories, along with motivational interviewing to ensure that content was tailored to individuals and their circumstances. The sessions and lesson plans were developed based on adult learning principles.

### Methods of assessment

Metabolic health outcome measures included body weight (in kilograms), waist circumference (in centimetres), Body Mass Index (BMI) and glycated haemoglobin (HbA1c). The HbA1c readings were collected by trained professionals drawing finger-prick blood samples and analysed using the Quo-Test HbA1c Analyzer. Changes in these metabolic measures were operationalised as *percentage change scores* to take into account variations in baseline and provide an optimal metric for characterising possible changes in outcome measures across interventions (see e.g., Cole et al., 2005; Hatoum & Kaplan, 2013). Alongside these a range of validated assessments relating to participant physical activity levels, diabetes and self-care behaviours, attitudes and beliefs, and quality of life assessments were completed by participants at each time point (see Table 1).

**Table 1.** Participant outcomes, psychometric scales and sources.

| Participant outcome | Psychometric scale | Reference |
| --- | --- | --- |
| Self-care and empowerment | Diabetes Empowerment Scale-Short Form (DES-SF) | Anderson et al. (2003) |
| Perceptions of diabetes | Revised Illness Perceptions Questionnaire (IPQ-R*) | Moss-Morris et al. (2002) |
| Eating behaviours | Dutch Eating Behaviour Questionnaire (DEBQ) | Van Strein et al. (1986) |
| Physical and mental health | Short Form Health Survey (SF-12) | Ware et al. (1996) |
| Self-care behaviours | Summary of Diabetes Self-Care Activities Measure (SDSCA) | Toobert et al. (2000) |
| Health-related quality of live | Visual Analogue Scale (EQ-VAS) from the European Quality of Life 5 Dimensions 3 Levels (EQ-5D-3L) | EuroQol (2009) |

\* **Note:** the brief version (BIPQ-R) was used in the Netherlands sample.

## Results

### Influence of DWELL programme on metabolic health outcomes

Percentage change scores in metabolic health were compared to zero using a one-sample Wilcoxon test as data were not normally distributed. Table 2 shows the median percentage change across the four metabolic measures from T0 to T1, and from T1 to T2, along with effect sizes (*r*) and indications of significance.

**Table 2.** Median percentage change scores for each metabolic measure from T0-T1 and T1-T2, with effect sizes and indications of significance.

| Metabolic measure | N | % Change T0-T1 | N | % Change T1-T2 |
| --- | --- | --- | --- | --- |
| Weight | 428 | -1.33% ($r=0.52$)*** | 252 | -0.09% ($r=0.15$)* |
| BMI | 424 | -1.31% ($r=0.48$)*** | 253 | -0.25% ($r=0.18$)** |
| Waist circumference | 406 | -1.70% ($r=0.47$)*** | 221 | 0.00% ($r=0.05$)ns |
| HbA1c | 380 | -4.59% ($r=0.39$)*** | 212 | 1.55% (0.16)* |

\* $p<0.05$ , \*\* $p<0.01$ , \*\*\* $p<0.001$

All percentage changes scores from baseline (T0) to end of programme (T1) were significantly greater than zero, indicating a clear reduction in all four metabolic measures immediately following the intervention. From end of programme (T1) to 6-month follow up (T2), percentage change scores for both weight and BMI were significantly greater than zero, showing clear reductions, while there was no further change of waist circumference while percentage change score for HbA1c showed a statistically significant increase by 1.55% from end of programme to the 6-month follow up.

There were strong positive correlations between the metabolic measure of *Weight* and *BMI*, and *Waist circumference* at the three time points, along with weaker positive correlations between *Weight* and *HbA1c* at T1 and T2 (see Table 3).

**Table 3.** Correlations between metabolic measure of Weight and BMI, Waist circumference and HbA1c at T0, T1 and T2.

|  | **T0** | **T1** | **T2** |
| --- | --- | --- | --- |
| BMI | 0.827*** | 0.729*** | 0.836*** |
| Waist circumference | 0.823*** | 0.816*** | 0.879*** |
| HbA1c | 0.020 | 0.113* | 0.134* |

\* $p < 0.05$ , \*\* $p < 0.01$ , \*\*\* $p < 0.001$

Given the relationship between *Weight* and the other metabolic measures, further exploration of possible associations between demographic variables and metabolic changes focused specifically on percentage weight loss change from T0 to T1.

### Influence of demographics on percentage weight loss

A linear regression analysis examined whether scores on the demographic measure of *Diagyears* (i.e., how many years since diagnosis) significantly predicted *Percentage Weight Loss Change*. There was a significant positive correlation between *Diagyears* and *Percentage Weight Loss Change* (*r*(336)=0.112, p<0.05) and whilst the model only explained 1.3% of the variance it was significant (*F*(1,334)=4.23, p<0.05), showing that *Diagyears* (*t* = 2.06, *p* < 0.05) was a significant predictor. No other demographic variables were a predictor of weight loss change.

### Influence of DWELL programme on psychological measures

Changes in assessed psychological measures from T0 (baseline) to T1 (post intervention) and from T1 to T2 (6 months follow-up) were examined using Wilcoxon tests as the data were not normally distributed. Findings are summarised in Table 4.

**Table 4.** Comparisons of median (SD) scores for psychological measures from T0 to T1 and from T1 to T2 with effect sizes and indications of significance.

| *Measure* | **T0** | **T1** | **T0 vs T1**<br>**Effect size** | **T2** | **T1 vs T2**<br>**Effect size** |
| --- | --- | --- | --- | --- | --- |
| Diabetes empowerment scale (DES-SF) |  |  |  |  |  |
| *Empowerment* | 28.0<br>(5.47) | 32.0<br>(4.58) | r=0.63*** | 31.0<br>(5.10) | r=0.15* |
| Positive perceptions of diabetes (IPQ-R) |  |  |  |  |  |
| *Illness coherence* | 15.0<br>(5.00) | 19.00<br>(5.25) | r=0.59*** | 19.00<br>(4.82) | r=0.02<sup>NS</sup> |
| *Treatment control* | 18.0<br>(4.31) | 19.00<br>(4.58) | r=0.14** | 19.00<br>(4.25) | r=0.14* |
| *Personal control* | 24.0<br>(6.26) | 24.00<br>(6.56) | r=0.29*** | 24.00<br>(6.29) | r=0.03<sup>NS</sup> |
| Negative perceptions of diabetes (IPQ-R) |  |  |  |  |  |
| *Timeline (acute/chronic)* | 23.00<br>(5.95) | 23.00<br>(6.53) | r=0.09<sup>NS</sup> | 24.00<br>(5.88) | r=0.12<sup>NS</sup> |
| *Consequences* | 19.00<br>(5.54) | 18.00<br>(5.42) | r=0.07<sup>NS</sup> | 18.00<br>(5.17) | r=0.04<sup>NS</sup> |
| *Timeline (cyclical)* | 12.00<br>(3.51) | 12.00<br>(3.62) | r=0.13* | 11.50<br>(3.46) | r=0.08<sup>NS</sup> |
| *Emotional* | 17.00<br>(6.52) | 16.00<br>(6.23) | r=0.35*** | 15.00<br>(6.02) | r=0.05<sup>NS</sup> |
| Eating behaviour (DEBQ) |  |  |  |  |  |
| *Restrained* | 29.00<br>(7.19) | 31.00<br>(8.33) | r=0.32*** | 30.00<br>(8.05) | r=0.22*** |
| *Emotional* | 34.00<br>(13.91) | 30.00<br>(13.89) | r=0.27*** | 28.00<br>(13.62) | r=0.04<sup>NS</sup> |
| *External* | 27.00<br>(7.28) | 25.00<br>(6.83) | r=0.26*** | 25.00<br>(6.74) | r=0.00<sup>NS</sup> |
| Health (SF12) |  |  |  |  |  |
| *Physical health* | 42.17<br>(10.90) | 46.14<br>(10.75) | r=0.22*** | 46.46<br>(10.75) | r=0.08<sup>NS</sup> |
| *Mental health* | 45.16<br>(10.84) | 48.85<br>(10.36) | r=0.25*** | 48.52<br>(10.13) | r=0.09<sup>NS</sup> |
| Self-care behaviours (SDSCA) |  |  |  |  |  |
| *Diet* | 2.00<br>(2.49) | 3.00<br>(2.56) | r=0.33*** | 7.00<br>(2.43) | r=0.76*** |
| Footcare behaviours (SDSCA) |  |  |  |  |  |
| *Washing* | 5.00<br>(2.25) | 6.00<br>(2.16) | r=0.16*** | 8.00<br>(1.50) | r=0.83*** |
| *Soaking* | 0.00<br>(1.98) | 0.00<br>(2.04) | r=0.05<sup>NS</sup> | 1.00<br>(2.71) | r=0.73*** |
| *Drying* | 3.00 | 5.00 | r=0.24*** | 8.00 | r=0.83*** |

|  | (2.57) | (2.40) |  | (1.97) |  |
| --- | --- | --- | --- | --- | --- |
| Adherence behaviours (SDSCA) |  |  |  |  |  |
| *Medication* | 6.00<br>(2.43) | 6.00<br>(2.25) | $r=0.10^*$ | 8.00<br>(1.09) | $r=0.85^{***}$ |
| *Insulin* | 9.00<br>(2.62) | 9.00<br>(2.42) | $r=0.04^{NS}$ | 9.00<br>(1.68) | $r=0.38^{***}$ |
| *Pills* | 6.00<br>(2.55) | 6.00<br>(2.48) | $r=0.22^{***}$ | 8.00<br>(1.30) | $r=0.78^{***}$ |
| Quality of life (EQ-VAS) |  |  |  |  |  |
| Quality of life | 70.00<br>(19.16) | 70.00<br>(19.56) | $r=0.30^{***}$ | 70.00<br>(17.14) | $r=0.07^{NS}$ |

\* $p<0.05$ , \*\* $p<0.01$ , \*\*\* $p<0.001$ , <sup>NS</sup> = not significant

As can be seen in Table 4, participants’ level of empowerment increased from T0 to T1, (N=414, Z = -12.823, p<0.001) and then ’plateaued’ at the 6-month follow up (N=255, Z = - 2.159, p<0.05).

Participants also showed a significant improvement of positive diabetes perceptions the end of the programme in terms of illness coherence, indicating better understanding of the condition (N=432, Z = -12.259, p<0.001), treatment control, indicating a stronger belief that treatment can control the condition (N=432, Z = -2.945, p<0.01) and personal control, denoting a stronger belief in personal control over diabetes (N=432, Z = -6.212, p<0.001). There was no further improvement at the 6-month follow up apart from treatment control (N=262, Z = -2.207, p<0.05). Also, improvements were noted in the negative perceptions of diabetes at the end of the programme; there was significant decrease in negative emotional responses (N=429, Z = -7.27, p<0.001) and perceptions that the illness has a cyclical nature (timeline - cyclical) (N=389, Z = -2.63, p<0.01), with a marginal reduction in perceptions that diabetes is short-term condition (timeline - acute/chronic) (N=432, Z = -1.93, p=0.054).There was no change in consequences, namely the perception that diabetes has significant negative impact (N=431, Z = -1.58, p=0.11). No further improvements were recorded in illness perceptions at the 6-month follow up.

With regards to eating behaviours, at the end of the programme, there was a significant increase in the level of restrained eating (N=409, Z = -6.41, p<0.001), and a decrease in both emotional eating (N=417, Z = -5.54, p<0.001) and eating in response to external food cues (N=420, Z = -5.28, p<0.001). At the follow up, there was a small decrease in the level of restrained eating (N=252, Z = -3.48, p<0.001) with no change in the other two types of eating behaviour.

Similarly, at the end of the programme, there were significant improvements in both physical (N=383, Z = -4.31, p<0.001) and mental health (N=383, Z = -4.90, p<0.001) with no further improvement at the 6-month follow up.

In terms of self-care behaviours, diet-related care - number of days spent spacing out carbohydrates - increased at the end of the programme (N=399, Z = -6.76, p<0.001) and continued to improve at the 6-month follow up (N=154, Z = -9.53, p<0.001). In relation to foot care, there were statistically significant improvements in washing (N=430, Z = -3.43, p<0.001) and drying feet (N=427, Z = -4.96, p<0.001) by end of the programme, with further improvement at the 6-month follow up in all footcare behaviours.

Importantly, participants reported at the end of the programme statistically significant improvements in medication adherence; in particular, adherence to prescribed medication (N=401, Z = -2.10, p<0.05) and pills (N=376, Z = -4.32, p<0.001), with no statistically significant change in the use of insulin (N=386, Z = -0.86, p=0.38). Improvements were sustained at the 6-month follow up.

Finally, improvements were reported by participants at the end of the programme in self-rated physical and mental health scores (respective means of 63.3 and 69.0; N=424, Z = -6.31, p<0.001). No further statistically significant improvement was noted at the 6-month follow up (N=148, Z = -0.90, p=0.36).

## Conclusions

Analysis of metabolic health and psychological outcomes for participants who completed the 12-week DWELL programme revealed improvements. Participants reported a significant reduction at the end of the programme in weight loss, BMI reduction, waist circumference reduction and HbA1c reduction. This change was maintained 6 months after the end of the programme (T2) in terms of weight loss and BMI. Length of diagnosis was a strong predictor of weight loss at the end of the programme and 6-month follow up.

Participation in the programme also led to enhanced levels of participant empowerment, with significant improvements also seen in perceptions of diabetes, eating behaviours, mental and physical health, and self-care behaviours. Most positive changes were maintained or even increased 6 months after the end of the programme.

Participants reported better understanding and more personal control over diabetes. They also felt better equipped to deal with life consequences associated with diabetes. Overall, participants reported reduced negative emotions associated with their diabetes and improved overall mental health.

Behaviours around eating and food also improved with better control and awareness of eating (restrained eating) and reduction in eating habits related to emotions (emotional eating) and external cues (external eating). Participation in the programme also led to improved adherence to dietary, footcare and medication advice. Broadly, participants reported receiving more advice on diet, exercise, blood sugar measurement and medication prescriptions after they had completed the programme. This might suggest that upon programme completion, having learnt more about diabetes, participants were more engaged with services, and were more receptive to specific diet advice, footcare and medication management. Wider cultural and healthcare systems factors could also have impacted on engagement with and uptake of services.

Analysis suggested that the effect of the DWELL programme reached a peak at the end of the programme delivery and follow up analysis highlighted broad patterns in sustained improvements in both metabolic and psychosocial outcomes; namely, in relation to weight loss; empowerment; treatment control; and self-care/footcare/medical adherence behaviours.

Overall, results suggest that the focus of the DWELL DSME programme on patient empowerment and self-control, by offering flexibility and choice of options as well as peer support, led to significant participant outcome improvements at the end of the programme delivery with sustained changes six months later in self-care behaviours, empowerment and weight loss. DWELL, a cross-European programme co-designed with patients, healthcare professionals and family carers, adds to the body of knowledge of patient-led DSME practice with a holistic approach. Further research could shed light on the cultural and intersectional aspects of such approach that can inform more targeted programmes supporting T2DM patients with multiple co-morbidities such as serious mental health conditions.

## Data Availability

All data produced in the present work are contained in the manuscript.

## Notes

### Competing Interest Statement

The authors have declared no competing interest.

### Funding Statement

The DWELL study was funded by the EU Interreg 2 Seas Mers Zeeen Programme 2014-2020 (co-funded by the European Regional Development Fund).

### Author Declarations

Health Research Authority/London Bridge Research Ethics Committee gave ethical approval for this work (REC reference: 17/LO/1480)

## References

Adu M.D., Malabu U.H., Malau-Aduli A.E.O. et al. (2019). Enablers and barriers to effective diabetes self-management: A multi-national investigation. PLoS One. 14(6), e0217771.

Anderson, R. M., Fitzgerald, J. T., Gruppen, L. D., et al. (2003). The Diabetes Empowerment Scale-Short Form (DES-SF). Diabetes Care, 26(5), 1641–1642.

Botha F, & Dahmann SC. (2023) Locus of control, self-control, and health outcomes. SSM Popul Health, Nov 24;25:101566. doi: 10.1016/j.ssmph.2023.101566.

Cadzow, R.B., Vest, B.M., Craig, M., Rowe, J.S. & Kahn, L.S. (2014). “Living well with Diabetes”: Evaluation of a Pilot Program to Promote Diabetes Prevention and Self-Management in a Medically Underserved Community. Diabetes Spectrum, 27(4), 246–55.

Captieux M., Pearce G., Parke H.L., et al (2018). Supported self-management for people with type 2 diabetes: a meta-review of quantitative systematic reviews. BMJ Open, 8, e024262.

Chowdhury HA, Harrison CL, Siddiquea BN, Tissera S, Afroz A, Ali L, Joham AE, Billah B. (2024). The effectiveness of diabetes self-management education intervention on glycaemic control and cardiometabolic risk in adults with type 2 diabetes in low- and middle-income countries: A systematic review and meta-analysis. PLoS One, 19(2), e0297328.

Cole, T. J., Faith, M. S., Pietrobelli, A., & Heo, M. (2005). What is the best measure of adiposity change in growing children: BMI, BMI%, BMI z-score or BMI centile? European Journal of Clinical Nutrition, 59(3), 419–425.

Craig P, Dieppe P, Macintyre S, Michie S, Nazareth I, & Petticrew M, (2008). Medical Research Council Guidance. Developing and evaluating complex interventions: the new Medical Research Council guidance. BMJ, 337, a1655.

Ernawati U, Wihastuti TA, & Utami YW (2021). Effectiveness of diabetes self-management education (DSME) in type 2 diabetes mellitus (T2DM) patients: Systematic literature review. J Public Health Res, 10(2), 2240.

EuroQol Group. (2009). EQ-5D-5L. Rotterdam: EuroQol Group. Available at https://euroqol.org/information-and-support/euroqol-instruments/eq-5d-3l/ (Accessed 24th June 2024).

Field, A. (2013). Discovering statistics using IBM SPSS statistics 4th Ed. London, Sage.

Hatoum, I. J., & Kaplan, L. M. (2013). Advantages of percent weight loss as a method of reporting weight loss after Roux‐en‐Y gastric bypass. Obesity, 21(8), 1519–1525. 10.1002/oby.20186

Hermanns, N., Ehrmann, D., Finke-Groene, K. & Kulzer, B. (2020). Trends in diabetes self-management education: where are we coming from and where are we going? A narrative review. Diabet. Med., 37, 436–447.

IDF. Diabetes Atlas (2021). International Diabetes Federation. 10th Edition. ISBN: 978-2-930229-98-0. Available at https://diabetesatlas.org/idfawp/resource-files/2021/07/IDF_Atlas_10th_Edition_2021.pdf (Accessed 13th July 2024)

International Diabetes Federation (2021). Diabetes now affects one in 10 adults worldwide.Available at https://idf.org/news/diabetes-now-affects-one-in-10-adults-worldwide/ (Accessed 13th July 2024).

Macaden, L. & Clark, C.L. (2010). The influence of locus of control on risk perception in older South Asian people with Type 2 diabetes in the UK. Nursing and Healthcare of Chronic Illness, 2, 144–152.

Moss-Morris, R., Weinman, J., Petrie, K., Horne, R., Cameron, L., & Buick, D. (2002). The Revised Illness Perception Questionnaire (IPQ-R). Psychology & Health, 17(1), 1–16.

Nam, S., Chesla, C., Stotts, N.A., Kroon, L., Janson, S.L. (2011). Barriers to diabetes management: patient and provider factors. Diabetes Research & Clinical Practice, 93(1), 1–9.

Nugent, L.E., Carson, M., Zammitt, N.N., Smith, G.D. & Wallston, K.A. (2015). Health value & perceived control over health: behavioural constructs to support Type 2 diabetes self-management in clinical practice. J Clin Nurs, 24, 2201–2210.

Riemenschneider H, Saha S, van den Broucke S, Maindal HT, Doyle G, Levin-Zamir D, Muller I, Ganahl K, Sørensen K, Chang P, Schillinger D, Schwarz PEH, Müller G. (2018). State of Diabetes Self-Management Education in the European Union Member States and Non-EU Countries: The Diabetes Literacy Project. J Diabetes Res., 17, 1467171.

Ribu L, Rønnevig M, & Corbin J. (2019). People with type 2 diabetes struggling for self-management: A part study from the randomized controlled trial in RENEWING HEALTH. Nurs Open, 6(3), 1088–1096.

Seuring, T., Archangelidi, O. & Suhrcke, M. (2015). The Economic Costs of Type 2 Diabetes: A Global Systematic Review. PharmacoEconomics, 33, 811–831

Sun H., Saeedi P., Karuranga S., et al (2022). IDF Diabetes Atlas: Global, regional and country-level diabetes prevalence estimates for 2021 and projections for 2045. Diabetes Res Clin Pract. 183, 109119. doi: 10.1016/j.diabres.2021.109119.

Toobert, D. J., Hampson, S. E., & Glasgow, R. E. (2000). The summary of diabetes self-care activities measure: results from 7 studies and a revised scale. Diabetes Care, 23(7), 943–950.

Vanbosseghem, R., Callens, A., & Luyens, V. (2020). DWELL Diabetes & Wellbeing. European Regional Development Fund.

Van Strien, T., Frijters, J. E., Bergers, G. P., & Defares, P. B. (1986). The Dutch Eating Behavior Questionnaire (DEBQ) for assessment of restrained, emotional, and external eating behavior. International Journal of Eating Disorders, 5(2), 295–315.

Ware, J. E., Kosinski, M., & Keller, S. D. (1996). A 12-Item Short-Form Health Survey: construction of scales and preliminary tests of reliability and validity. Medical Care, 34(3), 220–233.

Wilkinson, A., Whitehead, L. & Ritchie, L. (2014). Factors influencing the ability to self-manage diabetes for adults living with type 1 or 2 diabetes. International Journal of Nursing Studies, 51(1), 111–122.

Wilson V. (2021). Diabetes education to provide the necessary self-management skills. British Journal of Community Nursing, 26(4), 199–201.

World Health Organisation (online). Diabetes –Factsheet. Available at https://www.who.int/health-topics/diabetes#tab=tab_1 (Accessed 13th July 2024)

